# Clinical and Inflammatory Factors Influencing Constipation and Quality of Life in Cerebral Palsy

**DOI:** 10.1101/19011296

**Authors:** Ana Cristina Fernandes Maria Ferreira, Ryan J. Eveloff, Marcelo Freire, Maria Teresa Botti Rodrigues Santos

## Abstract

**Introduction:** Intestinal constipation is a clinical consequence, secondary to neuromotor disturbances, which acts on subjects with cerebral palsy (CP). The aim of this study was to investigate the factors influencing constipation and the quality of life (QOL) of CP subjects.

**Materials and methods:** We recruited a total of 63 subjects aging from 5-17 years with spastic CP who received physical rehabilitation. The subjects were divided into two groups including with and without constipation. Subjects were assigned into one of the 4 groups (G1-4) based on the prevalence of 1) CP and 2) Constipation. Subjects were assigned as CP with constipation (G1), CP without constipation (G2), and controls without CP with constipation (G3) and without CP and without constipation (G4). Subjects’ demographics, use of antiepileptic drugs (AEDs), motor function, caregiver priorities and child health index of life with disabilities (CPCHILD) were included. In addition to medical metadata, a subset of subjects was evaluated for oral and systemic inflammation through gingival bleeding and serum cytokine levels (TNF-α, IL-1β, IL-6, IL-8, IL-10) respectively. Statistical significance was evaluated by ANOVA One-Way (parametric data) and Kruskal Wallis (non-parametric data).

**Results:** A significant relationship was found between the type of medication and constipation. Subjects taking GABA and GABA+ (GABA in association with other medication) were more likely to be constipated than the other groups (P < 0.01). Additionally, quality of life was directly correlated with constipation; subjects in G1 presented the lower mean score of CPCHILD (49.0 #x00B1;13.1) compared to G2 (71.5 ± 16.7), when compared to G3 (88.9 ± 7.5), and G4 (95.5 ±5.0) (P < 0.01). Inflammation was more severe for patients in G1 (P < 0.001). There were no differences among groups regarding gender (P = 0.332) and age (P = 0.292).

**Conclusions:** Our results suggest that constipation was mostly affected by the use of certain antiepileptic drugs, namely GABA. This showed direct detrimental effect on CP quality of life, which was influenced by inflammatory cytokines and the dosage and type of AEDs.

## Introduction

Cerebral palsy (CP) is a life-limiting and costly disability [1]. CP is characterized by permanent neuromotor disorder affecting movement and by non-progressive degeneration of the brain [2]. As the major etiological factor for severe disability, the estimated prevalence of CP ranges from 2.3 to 2.9 per 1000 live births observed in the United States (2011–2012 National Survey of Children’s Health and the 2011–2013 National Health Interview Survey) [3]. Children with CP experience limitations in everyday activities, all of which are factors influencing their quality of life (QOL). It can be accompanied by difficulties related to perception, sensation, behavior, cognition, communication, and epilepsy. And, their QOL can be impacted by the development of musculoskeletal problems such as oral-gut motor impairment through muscle spasticity [2].

Due to an abnormal increase in muscle tone and injury to neural pathways, spasticity acts as a negative factor in the lives of 85-90% of individuals with CP [2,4,5]. The severity of movement disability is classified by Gross Motor Function Classification System (GMFCS) [6]. GMFCS is stratified into five levels of body mobility: Level I (walks without limitations), Level II (walks with limitations), Level III (walks using a hand-held mobility device), Level IV (self-mobility with limitations) and Level V (transported in a wheelchair) [6]. Our previous studies demonstrated that out of 254 subjects, 50 (19.7%) subjects had GMFCS I, II or III while 204 (80.3%) presented GMFCS IV or V [7–9]. These proportion results reflect on the population of this current cohort. It is worthy mentioning, that epilepsy prevalence increases in spastic CP subjects as the levels of GMFCS rise (IV and V) [10]. In fact, epilepsy affects 77% of the subjects with CP, and standard clinical treatment for epileptic CP patients is based on therapy with antiepileptic drugs (AEDs) [11]. As the principal inhibitory neurotransmitter of the central nervous system and also regulatory signal for muscle tone, Gamma Aminobutyric Acid (GABA) is used for neuronal excitability control (e.g.: benzodiazepines, phenobarbital, topiramate, vigabatrin, gabapentin enacarbil and ezogabine). Side effects of AEDs can range from subjective reports of mild drowsiness to life-threatening neurologic and gastrointestinal consequences [12].

The prevalence of severe constipation and lack of gastrointestinal control varies from 25% to 74% [3]. Constipation may be caused by multiple components including: 1) reduced intake of fiber and liquids (responsible for the digestive system functioning) [13], 2) central nervous system damage [14], 3) mobility reduction [15], 4) and/or the use of AEDs [13,16]. In subjects using AEDs the incidence of constipation is elevated [7]. Other side effects co-occurring with the gastrointestinal complications include oral dysbiosis, gingival bleeding (GB), and increase of systemic inflammation [17]. The exact clinical phenomic factors that are influenced by the use of AEDs and the impact on QOL remains elusive.

Here, we aimed to investigate the clinical factors that mostly influenced QOL in cerebral palsy subjects. Results demonstrated positive relationships of gastrointestinal constipation and the use of medication impacted the quality of life of CP patients. Of the AEDs tested, GABA was the strongest factor that influenced constipation, with significant results to gut and oral compartments. We further investigated the mechanism involved in the oral-gut phenotypes and assayed the inflammation as a catalyst for the clinical observations. For a subset of 37 participants, all of whom were CP subjects, we analyzed the salivary cytokines IL-6, IL-8, IL-10, IL1β, and TNF-α. Overall, the results demonstrated that, *via* elevated inflammation, the most constipated subjects had increased frequency of non-communicable diseases (oral and gut), leading to a decrease in QOL.

## Materials and Methods

### Study Design

This study was reviewed and approved by the Research Ethics Committee of the Cruzeiro do Sul University-Brazil Platform, São Paulo, Brazil (IRB #2,452,626). Written informed consent was obtained from the guardian of each child or adolescent after they were informed about the study. A cross-sectional study was performed with subjects with spastic CP diagnosis, who received a physical rehabilitation treatment at a referral center in São Paulo, Brazil, at the time of data collection.

### Participants

One hundred subjects with a medical diagnosis of CP were invited to participate in this study at AACD (Disabled Child Care Association) in São Paulo, Brazil. Inclusion criteria were a medical diagnosis of spastic CP raging from 5–17 years, both male and female, and presence or absence of constipation. Participants who presented progressive or neurodegenerative lesions or uncooperative behavior were excluded during clinical oral examinations. Data was collected from year of 2018 until 2019. Both demographic and clinical data were collected for each subject.

### Gastrointestinal (GI) Constipation

This study adopted the clinical constipation definition proposed by the Rome III criteria for constipation [18]. The individuals must present two or more of the following symptoms, in the previous three months, to be classified as presenting constipation including: (a) makes force to evacuate during at least 25% of defecations; (b) hard or irregular stools in at least 25% of defecations; (c) sensation of incomplete evacuation for at least 25% of defecations; (d) sensation of obstruction for at least 25% of defecations; (e) use of manual maneuvers in at least 25% of defecations; (f) less than three defecations per week.

The final sample of the study was composed of 30 subjects with CP with constipation (G1), 33 subjects with CP without constipation (G2), 7 subjects without CP with constipation (G3) and 23 subjects without CP and without constipation (G4). Data regarding gender, age, race (white, black and others), caregiver occupation and education, family income, medical diagnosis of CP according to the type of movement disorder (spastic), clinical pattern (tetraplegia, diplegia and hemiplegia), Gross Motor Function Classification System [6] (levels I to V) and use of AEDs were collected from their medical records. Caregivers answered the Caregiver Priorities and Child Health Index of Life with Disabilities [19] that consists of 36 items rated in six sections. CPCHILD is represented by the following domains: (1) Personal Care (eight items); (2) Positioning, Transfer, and Mobility (eight items); (3) Communication and Social Interaction (seven items); (4) Comfort, Emotions, and Behavior (nine items); (5) Health (three items); and (6) Overall Quality Of Life (one item).

### Saliva Collection

Unstimulated saliva samples were collected in dental assessment sessions. Subjects were asked to refrain from eating, drinking liquids or brushing their teeth for at least 1 hour prior to saliva collection. The collection was performed with the subjects sitting comfortably in a bright and ventilated room. Whole saliva was collected by passive flow for 5 minutes. After collection, the Salivette® was centrifuged at 5,000 rpm for five minutes at 4°C (Hettich Centrifuge, model Universal 320R, Tuttlingen, Germany) and frozen in a freezer at -80°C.

### Biomarker Sub Analysis

The analysis of cytokines in saliva was performed using a CBA Cytokine Inflammatory Kit (Becton Dickinson, CA, USA) for the detection of TNF-α, IL-1β, IL-6, IL-8, IL-10. All analyses were performed in duplicate. Briefly, 25μL of fluorescent particles conjugated to antibodies specific for each cytokine were added to 25μL of the saliva and incubated for one hour at room temperature away from light. Subsequently, 25μL of the secondary antibody conjugated to a fluorochrome was added to the mixture and incubated for two hours at room temperature. The results were compared to a standard curve with serially diluted cytokines. The particles were washed to remove unbound antibodies, resuspended in wash buffer and analyzed using a BD Accuri (BD Biosciences). Data acquisition was performed using BD-Accuri C6 Software, and concentrations were determined using FCAP software v.3.0 (BD Biosciences).

### Gingival Index (GI)

The evaluation of oral dysbiosis was assayed through the GI [20] by using a millimeter plastic periodontal probe (HuFriedy’s Colorvue PerioScreen Kit probe, Chicago, IL, USA), which was gently passed in the gingival margin of all teeth, in reference to the distobuccal papilla, the buccal margin, the mesiobuccal papilla and the lingual / palliative margin. Partially erupted teeth and residual roots were excluded without replacement. The index was calculated by the percentage of the sum of the individual values of each tooth divided by the number of faces examined. Classified as positive for gingivitis were the individuals that presented gingival marginal bleeding more than 20% of the total sites evaluated [20].

### Statistical Analyses

Analyses of descriptive statistics were performed to characterize the sample, calculate measures of central tendency and variability for the quantitative variables. The normality assumption of the quantitative variables was evaluated using the Shapiro-Wilks test. When normal distribution was observed, parametric tests were performed. Otherwise, non-parametric tests were selected to determine the significance of intergroups differences. The Chi-square test was used to analyze the variables of gender, presence of gingival bleeding, clinical type of CP, GMFCS, and AEDs used.

ANOVA One-Way (parametric data) was used to determine significant intergroups differences in relation to age and the periodontal condition studied. Kruskal Wallis(nonparametric data) was used to determine significant differences in relation intergroups cytokines levels. IBM SPSS Statistics (SPSS for Windows, Version 20.0, Armonk, NY: IBM Corp.) was used for all analyses, with a significance level at P<0.05. Data was visualized using Python 3.7 Anaconda with PyPlot and Seaborn and RStudio with ggplot2.

## Results

### 1 Participants Demographics

The goal of our study was to identify the main factors affecting QOL in CP subjects. We focused on first collecting subject information including demographics, clinical factors, and AEDs as potential influencers on a patient’s wellbeing and QOL (Table 1). In our population, 47 of our 93 subjects were female and 46 were male, accurately representing the relatively equal ratio of women to men. The sample power was calculated using means and standard deviations of overall domains among subjects with CP with constipation (51.0±13.1), subjects with CP without constipation (28.5±16.7), subjects without CP with constipation (11.1±7.5) and subjects without CP and without constipation (4.5±5.0) (OpenEpi online; www.openepi.com). The results showed that the G*Power at the 95% confidence interval was 96.88%.

**Table 1.** Population-Wide Demographics Table. Numbers and relevant statistics for study groups. Groups are defined as G1 (CP with constipation), G2 (CP without constipation), G3 (control with constipation), and G4 (control without constipation).

|  | G1 -<br>Constipated CP | G2-No<br>Constipated CP | G3- Constipated<br>Control <sup>a</sup> | G4- No Constipated<br>Control <sup>a</sup> | p - value * |
| --- | --- | --- | --- | --- | --- |
| **DEMOGRAPHICS** |  |  |  |  |  |
| Total Subjects | 30 (32.25%) | 33 (35.48%) | 7 (7.52%) | 23 (24.73%) |  |
| **Gender** |  |  |  |  |  |
| Female | 19 (66.3%) | 14 (42.4%) | 4 (57.1%) | 10 (43.5%) | 0.332 |
| Male | 11 (36.7%) | 19 (57.6%) | 3 (42.9%) | 13 (56.5%) |  |
| Age (mean±SD) | 8.5 ± 4.1 | 10.3 ± 3.7 | 8.8 ± 4.5 | 10.2 ± 4.3 | 0.291 |
| **Race** |  |  |  |  |  |
| White | 19 (63.4%) | 15(45.5%) | 6(85.7%) | 18(78.3%) |  |
| Others | 9(30%) | 16(48.5%) | 0(0%) | 5(21.7%) | 0.08 |
| Black | 2(6.4%) | 2(6%) | 1(14.3%) | 0(0%) |  |
| **Occupation (Caregiver)** |  |  |  |  |  |
| Employed | 11 (36,66%) | 13 (39,39%) | 5 (71,42%) | 19 (82,60) | 0.0017* |
| **Education** |  |  |  |  |  |
| ≤ 8 years | 21 (70%) | 23 (69.69%) | 4 (57.15%) | 14 (60.86%) | 0.5628 |
| > 8 years | 9 (30%) | 10 (30.31%) | 3 (42.85%) | 9 (39.14%) |  |
| **Family (Income)** |  |  |  |  |  |
| ≥3 MW | 4 (13.34%) | 9 (27.28%) | 3 (42.86%) | 11 (47.82%) | 0.018* |
| <3 MW | 26 (86.66%) | 24 (72.72%) | 4 (57.14%) | 12 (52.17%) |  |
| **ORAL HEALTH- PERIODONTAL PARAMETERS** |  |  |  |  |  |
| Gingival Bleeding | 32.4 ± 19.8 | 4.1 ± 3.5 | 0.6 ±1.4 | 0.1 ± 0.6 | <0.001* |
| **Gingivitis** |  |  |  |  |  |
| Presence | 23 (76.7%) | 2 (6.1%) | 0 (0%) | 0 (0%) | <0.001* |
| Absence | 7 (23.3%) | 31 (93.9%) | 7 (100%) | 23 (100%) |  |
| **SYSTEMIC HEALTH- CEREBRAL PALSY** |  |  |  |  |  |
| **Clinical Pattern** |  |  |  |  |  |
| Tetraplegia | 20 (66.66%) | 9 (27.27%) | 0 (0%) | 0 (0%) | 0.0009* |
| Diplegia | 10 (33.33%) | 15 (45.45%) | 0 (0%) | 0 (0%) |  |
| Hemiplegia | 0 (0%) | 9 (27.27%) | 0 (0%) | 0 (0%) |  |
| **GMFCS** |  |  |  |  |  |
| I - II - III | 0 (0%) | 14 (42.4%) | 7 (100%) | 23 (100%) | 0.0002* |
| IV - V | 30 (100%) | 19 (57.6%) | 0 (0%) | 0 (0%) |  |
| **Medication** |  |  |  |  |  |
| GABA | 5 (16.7%) | 1 (3%) | 0 (0%) | 0 (0%) |  |
| GABA + | 9 (30%) | 2 (6.1%) | 0 (0%) | 0 (0%) |  |
| Sodium | 5 (16.7%) | 4 (12.1%) | 0 (0%) | 0 (0%) | 0.0063* |
| Calcium | 1 (3.3%) | 1 (3%) | 0 (0%) | 0 (0%) |  |
| No medication | 10 (33.3%) | 25 (75.8) | 7 (100%) | 23 (100%) |  |
| **Quality of Life** |  |  |  |  |  |
| Personal Care | 79.8 ± 21.6A | 46.4 ± 37.5B | 20.0 ±23.8C | 3.8 ± 8.4C | <0.05* |
| Positioning, Transfer ar | 70.7 ± 26.1A | 42.1 ± 34.4B | 0.0 ±0.0C | 0.1 ± 0.5C | <0.05* |
| Communication and So | 44.4 ± 28.3A | 17.9 ± 19.3B | 0.0 ±0.0C | 0.0 ±0.0C | <0.05* |
| Comfort, Emotions and | 40.6 ± 22.7A | 18.8 ± 16.00B | 8.7 ±12.8C | 3.9 ± 4.9C | <0.05* |
| Health | 26.4 ± 18.2A | 16.2 ± 12.9B | 9.3±10.2C | 3.5 ± 5.7C | <0.05* |
| Overall Quality of Life | 44.3 ± 24.3A | 29.7± 15.2B | 28.6 ±19.5C | 18.3 ± 20.8C | <0.05* |
| Domains Means | 51.0 ± 13.1A | 28.5 ±16.7B | 11.1 ±7.5C | 4.5 ±5.0C | <0.05* |

<sup>a</sup> Values represent mean ± SD or percentage (%); \* p-values for group comparisons were significant at 0.05; Chi-square, ANOVA, One-Way, Kruskal-Wallis tests. Different letters denote statistically significant differences (p<0.05). Uppercase letters compare values horizontally (intragroup evaluation).
*Abbreviations* : SD-standard deviation; BOP-Bleeding on probing; G1-CP with constipation; G2-CP without constipation; G3- without CP with constipation; G4- without CP without constipation; MW- minimum wage, unit for measuring income in Brazil, broadly corresponding to \$251 U.S. during the study period

There were no significant differences among groups regarding gender (P = 0.332), age (P = 0.291), race (P = 0.08), and education (P = 0.5628), yet significant differences were found on caregivers occupation (P = 0.0017) and family income (P = 0.018). G1 was composed of 30 non-ambulatory subjects (GMFCS levels IV and V) and G2 of 14 ambulatory subjects (GMFCS levels I, II, and III) (P = 0.0002). Worse QOL measures were observed for constipated subjects presenting the clinical pattern tetraplegia when compared to non-constipated CP (P < 0.001) (Table 1). Interestingly, gender was significantly associated with constipation (P < 0.05) with females appearing more likely to be constipated in our population. Distribution of age in our population shows a peak at ∼6 years of age, with a slight steady dropoff in population representation until approximately age 14, with a steep dropoff in population representation from ages 14-19 (Figure 1A).

**Figure 1.**
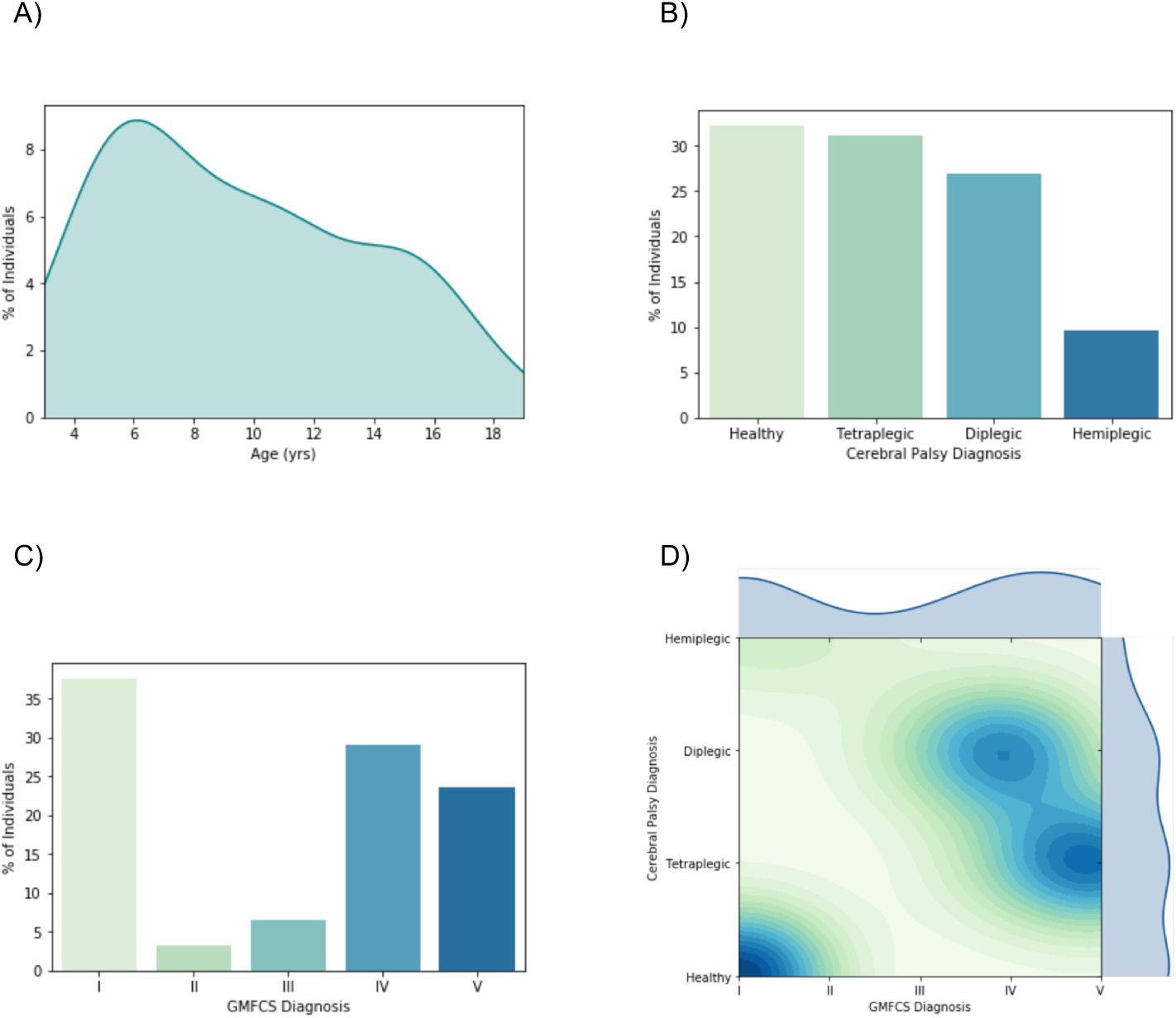
Distribution patterns of the CP study population. **A)** Subjects from the cohort aged from 3-19, with a mean age of 9.57 ± 4.20 years (n=93). **B)** The study population was composed of healthy individuals (controls, 32.26%), tetraplegics (32.26%), diplegics (26.88%), and hemiplegics (9.68%). **C)** Distribution is measured along the GMFCS scale from I-V. Stages I-III are considered ambulatory, while stages IV and V require significant assistance (typically a wheelchair or mobility scooter). Stages II and III are less represented (3.23% and 6.45% respectively), while stages I (37.63%), IV (29.03%) and V (23.66%) make up the vast majority of our population. **D)** Clustering shows that hemiplegics and healthy individuals are more likely to be ambulatory, most without assistance. However, diplegic and tetraplegic individuals are by definition not ambulatory, due to the increased muscle tone. Therefore, diplegics are frequently classified as GMFCS stage IV and tetraplegics as stage V.

### 2 CP Diagnosis

When diagnosing CP, there are three widely accepted presentations: 1) hemiplegic, or Increased muscle tone in a hemibody; 2) diplegic, Increased muscle tone in lower limbs (may affect upper limbs, but to a lesser extent); and 3) tetraplegic, a system-wide decay of neuromotor function, represented by increased muscle tone in all four limbs and trunk involvement. In our population, the distribution of subjects is made up of these three groups as well as healthy individuals (control). Hemiplegia is the least prominent in our population, with 9.68% of subjects presenting hemiplegia, 26.88% diplegia, and 31.18% tetraplegia (32.26% of our population was healthy at examination) (Figure 1B). CP diagnosis was found to be significant in our population (P < 0.001) in determining QOL (Table 1).

Another mechanism to classify subjects with spastic CP is according to the motor function ability. The Gross Motor Function Classification System (GMFCS classify as ambulatory that ones who present levels I-III, while levels IV and V are reserved for non-ambulatory subjects. In our population, levels I (37.63%), IV (29.03%) and V (23.66%) make up the vast majority of our population (Figure 1C). Level I is mostly populated by our control group, while groups IV and V are reserved for subjects with severe CP, namely diplegic and tetraplegic CP (Figure 1D). More specifically, group IV is made up of diplegics subjects who can typically operate a wheelchair manually, while tetraplegic subjects require caregiver-assisted or motorized forms of transportation.

We found that a subject’s GMFCS level was statistically significant in determining their QOL (P < 0.001). Functional motor independence of subjects with CP is directly related to the extent of cortical injury. Diplegic and quadriplegic subjects (GMFCS IV and V) have significant motor impairment that limits daily living activities. Therefore, the diagnosis of CP and greater motor damage are two strong factors that influence the quality of life of the subjects.

### 3 Constipation Impact on Quality of Life

According to CPCHILD, the groups differed significantly (P < 0.05). G1 presented higher values (lower QOL) for all sections of the questionnaire while compared to G2, G3 and G4. The CPCHILD questionnaire measures QOL presented 6 subscores: 1) Positioning, Transferring, and Mobility; 2) Caregiver-Reported QOL; 3) Hospitalization and Caregiver-Reported Health; 4) Comfort and Emotions; 5) Personal Care; and 6) Communication and Social Interaction. These subscores are scaled from 0 to 100 and were averaged to determine overall QOL. The combination of constipation and cerebral palsy, showed to lower the quality of life of each individual. The inability to effectively and routinely have bowel movements and the inhibition or even incapability to move freely leads to fewer opportunities and consequently lower overall QOL.

In our initial analysis, we uncovered the aspects of subject’s life that determine their overall QOL. In figure 2, each subscore is plotted in relation to the remaining 5 subscores. For example, figures 2A-F show Hospitalization and Caregiver-Reported Health (HCRH) compared to all other subscores. A striking trend is observed wherein a particular subscore (in this case) was not particularly influential in determining overall QOL, as a rating of 80 in HCRH can still yield QOL scores as low as 20. Conversely, Communication and Social Interaction (CSI) scores much more closely reflect QOL scores as a whole. Gender was not significantly correlated with QOL in our population. Further analysis into what physical and psychological factors that most directly pertain to a subject’s QOL is required to better understand and treat CP subjects.

**Figure 2.**
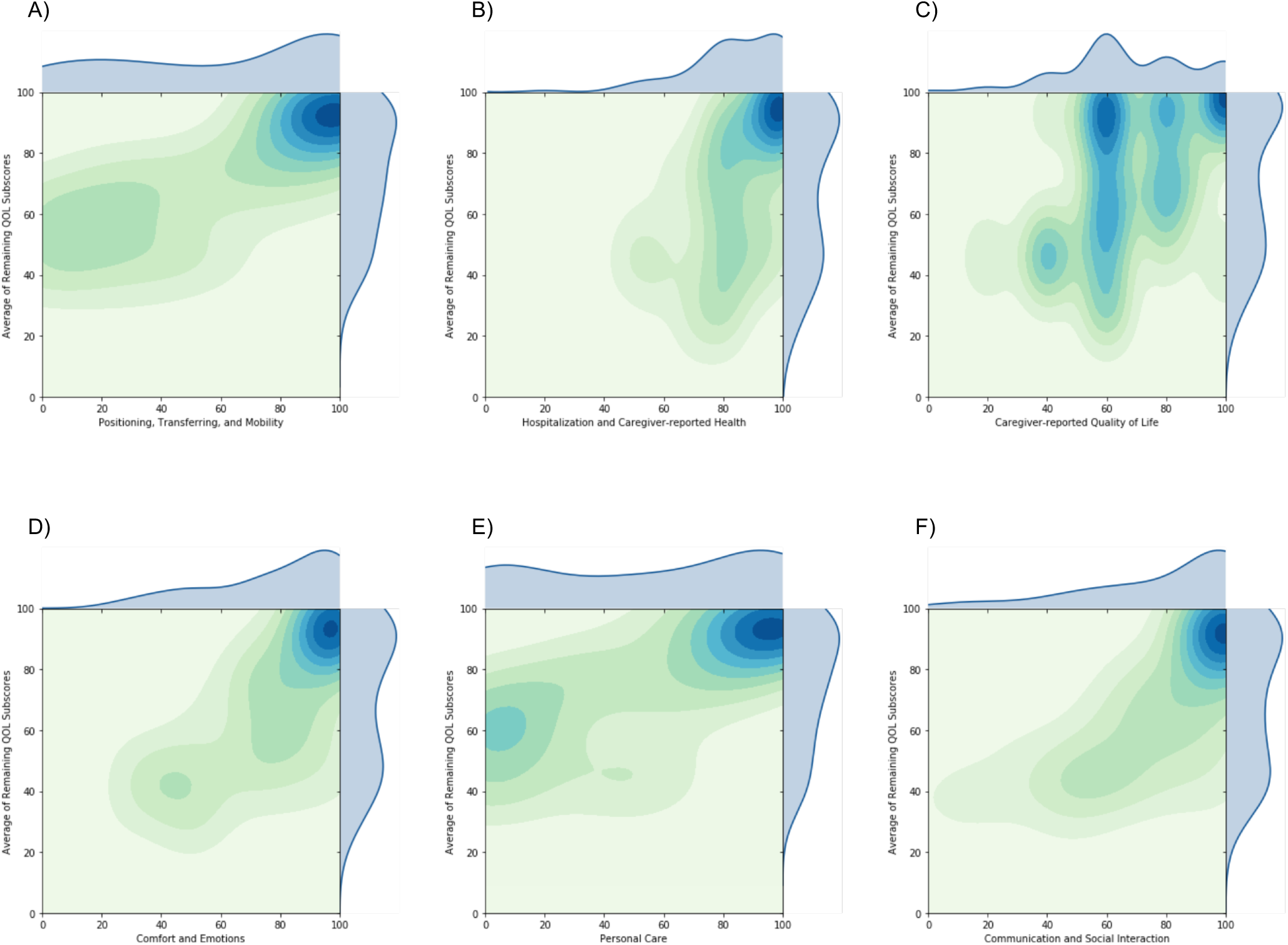
Effects of CPCHILD Subscores on Overall QOL. Distributions of each individual CPCHILD subscore A) Positioning, Transferring, and Mobility; B) Hospitalization and Caregiver-Reported Health; C) Caregiver-Reported Quality of Life; D) Comfort and Emotions; E) Personal Care; and F) Communication and Social Interaction) is measured on the x-axes, while the y-axis marks the average of the remaining subscores. A lighter green represents a lower density within the distribution, while a darker blue represents a higher density.

### 3 Medication Affecting Constipation

The key factor affecting QOL and constipation in our population was revealed to be medication types. As the common thread linking all CP subjects, pharmacological treatment is an essential point of emphasis of our study. Subjects that use of GABA (the principal inhibitory neurotransmitter in the central nervous system and regulate muscle tone) and GABA+ presented the worst quality of life. The use of sodium inhibitors (Na inhibitor) were positively correlated with a higher quality of life, potentially due to the removal of sodium from the digestive tract, yielding softer stool (Figure 1 A-C). Specifically, the choice of AED used for treatment of CP and epilepsy greatly influenced the physical and mental health of our subjects. GB is seen in figures 3D-F to be higher in subjects taking GABA and GABA+, or GABA in association with other AEDs. Conversely, subjects taking sodium inhibitors were clustered at lower mean GB.

**Figure 3.**
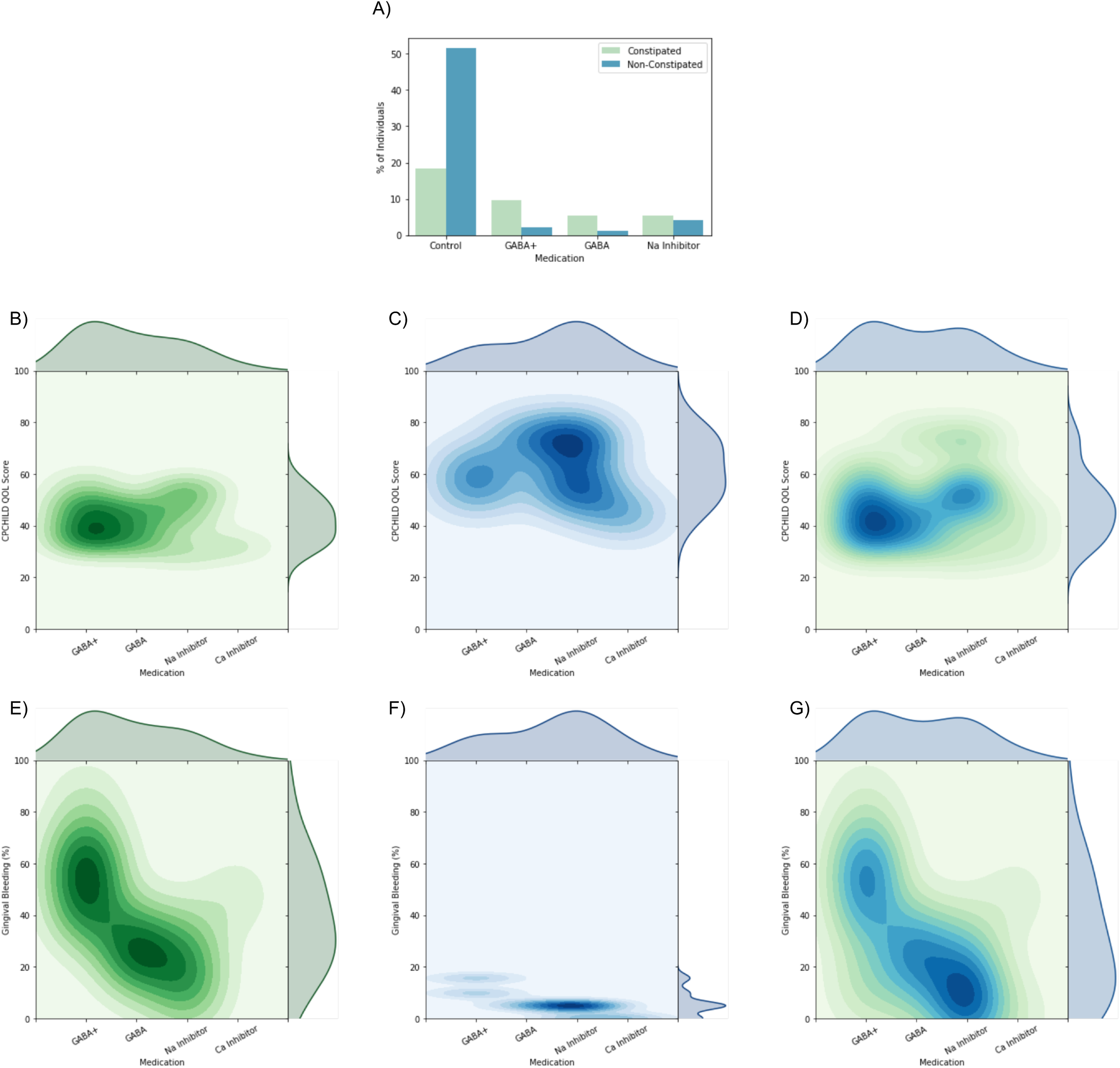
Correlation of Clinical Factors in CP patients. **A)** Each KDE plot shows clustering by the type of medication taken by subjects. Green figures show constipated subjects, blue figures show non-constipated subjects, and Green-to-Blue figures show the full population. Constipation is measured and compared to the medication types. In **B-D)** QOLS is scored from 0-100 based on the average of CPCHILD subscores. **E-G)** gingival bleeding is measured by the percentage of teeth after periodontal probing.

Sodium inhibitors were associated with a higher QOL and GABA (the chief inhibitory neurotransmitter in our body) was associated with a lower QOL. Similarly, GABA+ was associated with a significantly lower QOL. In Figure 3G, there is a clear trend wherein GABA subjects are clustered at a lower QOL, while sodium inhibitor subjects, particularly in our non-constipated subcohort (Figure 3H), exhibit a clustering at a much higher QOL. Calcium inhibitors were only taken by 2 subjects in our population, so further studies must be performed with larger populations to fully understand their effect on the QOL and bowel health of CP and epilepsy subjects.With this information, it is clear that the adverse effects of GABA on a subjects’ body and mental wellbeing must be observed further; similarly, sodium inhibitors should be studied to determine their efficacy as an AED as a replacement for or synergistically with GABA.

### 4 Oral and Systemic Inflammation are Elevated in Constipated Patients

From a clinical perspective, CP is considered a neurological disease. However, its symptoms can manifest themselves throughout the body. In many subjects, the lack of an ability to care for oneself leads to the development of oral and gum disease. These diseases manifest themselves through bleeding of the gingiva. If a large number of teeth have bleeding gingiva, there is a high chance of oral disease manifesting itself as gingival inflammation and periodontitis.

In figure 4A, Density for males (green) appears more concentrated at lower levels of Gingival bleeding (GB). Females (blue) are more stratified, but still show a higher density where GB is < 20%. The average GB observed for males was 8.37±12.18%, while the same for females was 16.70±22.26%. Overall mean GB was 12.08±18.27%. In figure 4B, a slight visual trend is observed with the upper limit of mean GB decreasing with age. Higher densities of high gingival index occur in younger subjects, while higher densities of low gingival bleeding occur in older subjects. Healthy individuals (those who do not suffer from CP) have markedly lower mean GB (23%) than any other group, with 2.79% mean hemiplegic GB, 12.19% mean diplegic GB, and 27.11% mean tetraplegic GB (Figure 4C).

**Figure 4.**
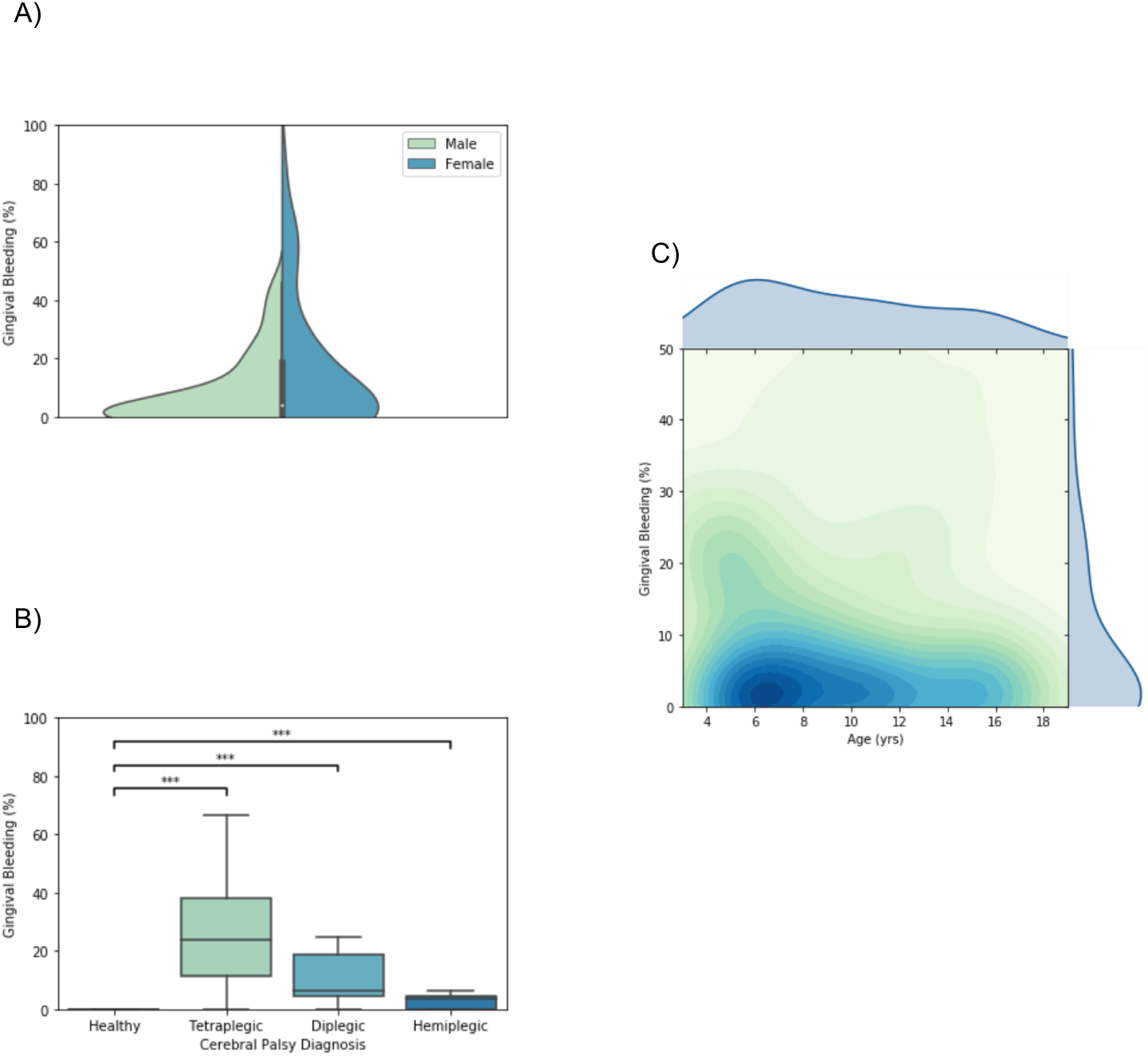
Factors impacting gingival bleeding. **A)** Violin plots depicting gingival bleeding patterns stratified by gender. Box plot within the violin represent the inner quartiles, and the whiskers show the 5% confidence interval (CI). KDE clustering demonstrating gingival bleeding in relation to age distribution. Data represented as means ± sd, and significant level at p-value < 0.05 (n=93). **C)** Gingival bleeding according to CP diagnosis. Boxes represent inner quartiles, center line represents the mean, while whiskers represent 5% CI (***, *P* < *0*.*001*).

Indirect symptoms of CP are not limited to the oral cavity; gastrointestinal problems can often arise as a reaction to medications or from an inability to properly digest and pass food. Of our 63 subjects who present CP, 30 were constipated at examination, representing a drastically higher prevalence of constipation (47.62%) compared to the estimated prevalence of 16% in the global population [21].

For a subset of our population, we collected salivary cytokine measures to get an accurate idea of systemic inflammation and its relationship with constipation and medication. For all cytokines for which data was collected (IL-6, IL-8, IL-10, IL-1β, and TNF-α), mean levels approximately doubled in constipated subjects. In figures 5A-E, there is a clear visual trend of higher resting systemic inflammation in individuals with severe CP. Of these cytokines, only IL1β was significantly correlated with constipation (P < 0.05, Figure 5D).

**Figure 5.**
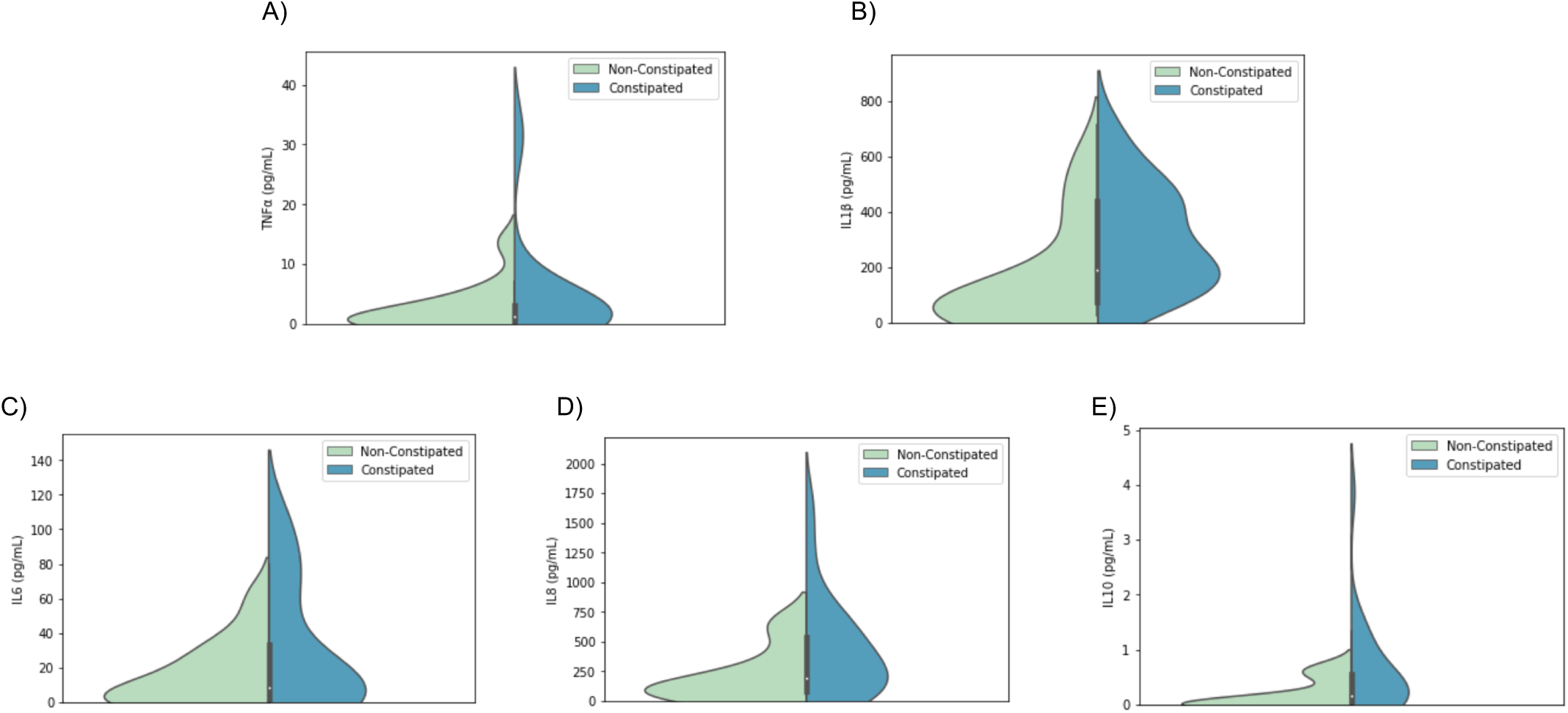
Constipation actions on Inflammatory Cytokine Levels. Distributions of each measured cytokine [A) TNF-α, B) IL-1β, C) IL-6, D) IL-8, and E) IL-10] in constipated subjects (blue) and non-constipated subjects (orange) are mapped using a violin plot. White dots mark the means, bars show the inner quartiles, and whiskers mark the 5% confidence interval.

Figure 6 reveals the positive and negative correlations in our study cohort among clinical factors and demographics. Naturally, each subscore is correlated with the overall QOL of an individual as they are part of its calculation. Key trends to notice are: 1) The presence of constipation significantly lowers QOL; 2) GMFCS and CP Type are strongly correlated, both to one another and to QOL; 3) GB exhibits a strong negative correlation with QOL that is statistically significant; and 4) Overall QOL is more strongly correlated to the self-reported mental health of -subject than to caregiver-reported metrics.

**Figure 6.**
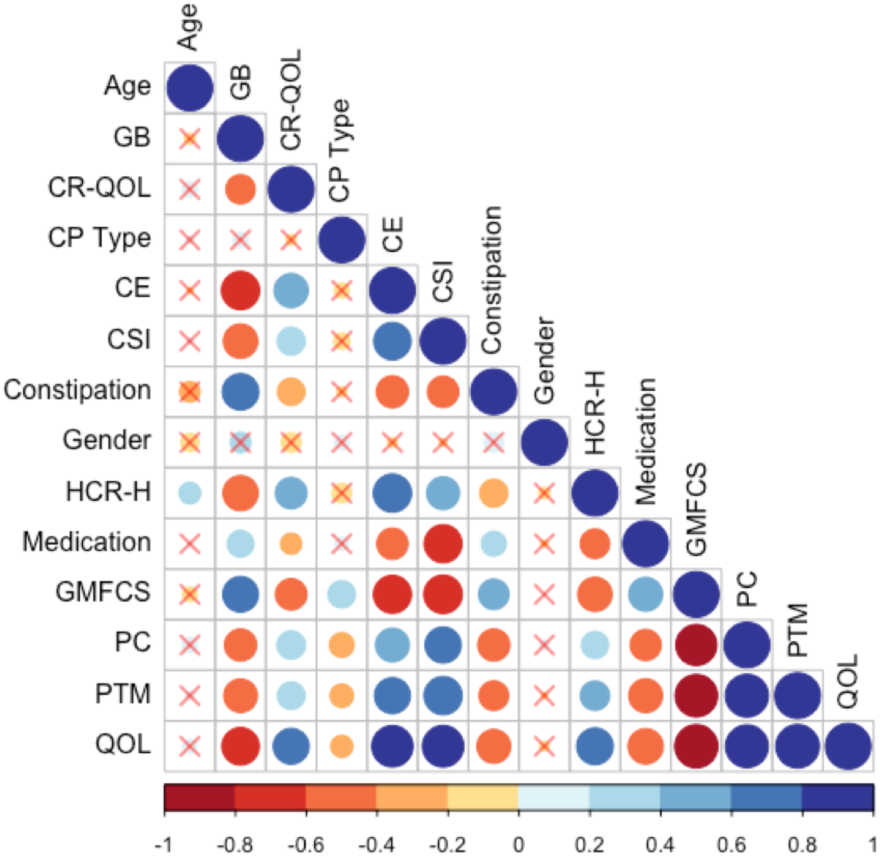
Correlation of Clinical Factors in CP patients. Correlogram of Pearson’s correlation of clinical factors influencing CP patients. Each axis is labeled by relevant factors to form a lower-triangle correlogram. Each dot represents a significant correlation coefficient of two factors assigned (P < 0.05), where size and saturation represent the magnitude of the correlation, and the color itself represents whether the correlation is positive (blue) or negative (red), following the scale bar. Dots are overlaid by an X represent associations and are not significant regardless of the strength. (GB–Gingival Bleeding, CR-QOL–Caregiver-Reported Quality of Life, CE–Comfort and Emotions, CSI–Communication and Social Interaction, HCR-H–Hospitalization and Caregiver-Reported Health, GMFCS–Movement Ability, PC–Personal Care, PTM–Positioning, Transferring, and Mobility).

## Discussion

The results provided novel information related to the severity of CP and the consequences of treatment on constipation, oral health, and quality of life. To the best of our knowledge, this is the first study to evaluate the association between intestinal constipation and quality of life of CP using the Caregiver Priorities and Child Health Index of Life with Disabilities. According to the motor type disorder, subjects with CP are classified as at least one of spastic, dyskinetic (dystonic, choreic and athetosic), or ataxic. The subjects presented spastic CP, the most prevalent type of motor disorder, with tetraplegia the clinical pattern most prevalent (32.26%) (Figure 1B). Consistent with our results, tetraplegia was also observed in a 3,294 children study developed in 18 states in the USA (32.9%) [22]. Early brain damage triggers complex adaptive neuroplasticity processes involving multiple functional systems. Different factors interact with brain plasticity potentials, influencing the natural history of cerebral palsy in the early years of childhood [23]. The later the interdisciplinary intervention, the smaller the gain during brain plasticity, reducing the potential for improvement in the individual’s condition.

Here, we observed a shortage of national instruments, which appears to reflect the difficulty of the scientific community to develop tools for assessing quality of life that apply to the socio-cultural diversity of the country. For all questionnaires revised, only Caregiver Priorities and Child Health Index of Life with Disabilities (CPCHILD) [19] and Cerebral Palsy Quality of Life Questionnaire for Children Child report (CPQol-Child) [24] are specific to Cerebral Palsy subjects. CPQOL-Child is the individual’s own report, since it was mostly used in children with GMFCS level I or II and were excluded the individual with any intellectual deficits and lack of an efficient communication system. In this study, the largest number of individuals belonged to GMFCS level IV and V (Figure 1C) and due to the difficulty of communication, their caregivers answered the questionnaire as their proxy; in this case the CPCHILD is the best choice of questionnaire. The other questionnaires were for children and adapted for a wide range of disabilities, while CPCHILD is more specific to our study population. The Cronbach’s alpha value estimated for the Caregiver Priorities and Child Health Index of Life with Disabilities was 0.879. It was observed that G1 presented a higher mean score of CPCHILD (51.0 ±13.1) compared to G2 (28.5 ± 16.7), G3 (11.1 ± 7.5), and G4 (4.5 ±5.0) (P < 0.05) (Table 1).Seventy percent of caregivers of CP subjects in this study had less than 8 years of study (Table1). Intellectual capital, represented by education, increases the likelihood of access to information, health-related issues [25] facilitating understanding and recognizing the importance of self-care. A low level of caregiver education influences the child’s overall health, represented by self-care of general and oral health [26].

The possible explanation for the lower education level of caregivers for individuals with CP is the ‘caregiver martyr syndrome’ wherein a caregiver neglects their own health, studies, and finances to provide more frequent and consistent [27] care [27]. With a lower educational level and fewer paid employment opportunities, there is a direct logical connection linking significantly lower income to caregivers of individuals with CP (Table 1). Caregivers’ potentially lower standard of education prevents them from entering the skilled labor market. Thus, they dedicate themselves exclusively to household chores and taking care of the individual with CP [28]. Emerging concepts of quality of life related to CP are emerging, and in the last five years, a total of 469 articles were found in the literature.

We noticed that the severity of pain and discomfort has been associated with increased motor impairment and other comorbidities, particularly constipation and spasticity. In individuals with minor GMFCS, pain was observed during voluntary movements, whereas in individuals with larger GMFCS, pain was directly related to movement performed by the caregiver or therapist [29]. Higher intensity of pain and discomfort had a negative effect on physical and psychological quality of life (Figure 2A–F). Communication and social interactions in individuals with cerebral palsy in GMFCS level V with intellectual disability develop less favorably and show large variation [30] (Figure 2F). Fairhust et al. concluded in 2018 that increasing awareness of CP pain and comorbidities, particularly overall health and constipation, can help prevent and treat the disorder more effectively, increasing patients’ overall quality of life.

Medication such as AEDs are frequently used for the control of epileptic seizures [31]. These drugs can be administered in monotherapy or polytherapy, depending on the response to seizure control [32]. Results demonstrated that individuals with a lower quality of life were positively correlated to those who use polytherapeutic GABA treatments (GABA+), while those who use monotherapy, especially sodium channel inhibitors, have a better quality of life (Figure 3C) when compared with GABA and GABA+ (Figure 3B). AEDs have an effect on voltage-dependent modulation of sodium, calcium, and potassium channels, and the Aminobutyric Gamma Acid (GABA) is considered the primary inhibitory neurotransmitter not only in the central nervous system, but also on enteric nerves [32]. However, AEDs aims to achieve seizure control are usually limited by toxicity and adverse effects that impair an individual’s quality of life [33] (Figure 3B-D). The subjects taking GABA or GABA association presented a negative correlation with quality of life and also present intestinal constipation not only for the inhibitory GABA action, [34] but may also be related to dysbiosis.

A higher prevalence of gingivitis was observed in constipated subjects from this study, that were in use of GABA/or GABA+ (Figure 3E-G, *Supplementary Figure 1*). G1 presented significantly higher percentages of subjects with gingivitis compared to all other groups (P < 0.001). Additionally, CP subjects using GABA or GABA+ who were constipated had higher levels of gingival bleeding than those who did not (P = 0.006, Table 1). Current literature indicates that intestinal disorders play a prominent role in inflammatory responses to neurological conditions [35]. This line of evidence is fundamental to identifying the effects of dysbiosis on mucosal inflammation throughout the digestive tract. Significantly higher levels of IL1β, IL6, IL8, and IL10 were found in constipated subjects with gingival bleeding from this study (Figure 5), indicating an ongoing inflammatory process and the progression of periodontal disease [36–40]. Since the use of these medications cause CP subjects to present a reduced salivary flow rate, an increased value of salivary osmolality, dry mouth and gingivitis, which is represented by higher levels of inflammatory cytokines in quadriplegics [17]. Sodium inhibitors, however, have a less severe correlation with gingival bleeding (R^2^=.684), but have markedly lower effects on a patient’s QOL and they are associated with lower levels of constipation, it can be assumed that sodium inhibitors carry less of a systemic reaction in our study population.

Constipation is a common comorbidity described in CP subjects [13], and current literature agrees that constipation is the causal factor of gut dysbiosis [41,42]. Constipation prevalence observed in this study was higher (47.62%), with higher prevalence in females CP subjects (Figure 4). Females without CP present constipation and almost 2 times more likely to suffer from constipation than males [43]. Constipation and intestinal dysbiosis leads to increased mucosal permeability (leaky-gut) [42] of the Gut-Brain Axis [44] increasing serum endotoxin concentration. These endotoxins activate the immune system and promote IL1β production as observed in individuals with Autism Syndrome Disorder [45]. Moreover, there has been no published research to date that evaluated the effect of dysbiosis on gingivitis development in individuals with cerebral palsy.

In addition to GI and oral inflammation, we have observed gender to be an important variable in regard to inflammation. Females presented higher pro-inflammatory cytokine expression when compared to males (Figure 4). This gender bias has not been addressed in the literature and will require further studies. The possible explanation for these findings may be related to the phenomenon that CP occurs more frequently in male [46] categorized as GMFCS V [47]. Due to this limitation, these subjects require more advanced care, notably oral care and hygiene performed by the caregiver. Perhaps the females were able to perform oral hygiene on their own due to fewer neuromotor limitations, which in turn may have resulted in greater gingival bleeding. Additionally, hormonal changes in puberty should be examined as a factor for this observed gender dichotomy [48].

Inflammatory biomarkers can be evaluated both in serum [49] or in saliva [8]. Blood collection in subjects with CP is a hard task, because these subjects present sympathetic nervous system predominance [50], resulting in vasoconstriction, and consequently making peripheral venous access more difficult. On the other hand, saliva is readily available and its components are easy to be collected, and that has been used to evaluate salivary parameters in subjects with CP [17]. Ideal for future longitudinal monitoring.

The comparison between the subjects constipated and non-constipated from this study demonstrated that constipation acts as a damaging factor causing approximately doubled mean cytokine production (Figures 5A-E). In the presence of dysbiosis, virulence factors are released from pathogenic microorganisms present in the oral cavity, activating host’s immune-inflammatory responses [51–53]. IL1-β is an inflammatory cytokine associated with innate immune response, inflammation, pathogenesis and progression of periodontal disease [54,55]. Higher levels of IL1β (P < 0.05) were found in this study’s subjects, who were constipated and presented higher values of gingival bleeding (Figure 5B, *Supplementary Figure 2B*). Previous studies showed higher levels of IL-1β IL-6, IL-8 in spastic CP subjects presenting GMCFS level V [17].

Significantly higher levels of IL-6, IL-8 and IL-10 were found in constipated subjects with gingival bleeding (Figure 5C-E; *Supplementary Figure 2C-E*). These cytokines represent the action of pro-inflammatory and anti-inflammatory cytokines [55], critical biomarkers of periodontal inflammation corresponding to the clinical severity of the disease [37]. IL-6 and IL-8 were also described as potential predictors for oral diseases, reinforcing the importance of evaluating the salivary levels of these biomarkers [56]. The presence of the chemokine IL-8 induces the secretion of lymphocytes, monocytes, epithelial cells, fibroblasts, tumor cells, bone resorption and IL-1β, indicating an ongoing inflammatory process and the progression of periodontal disease [38,39]. IL-10 acts as an anti-inflammatory cytokine, inhibiting pro-inflammatory cytokines IL-1 and IL-6 associated with improvements in periodontal clinical parameters [57]. However, the degree of mucosal inflammation of constipated CP subjects represented by gingival bleeding was so high that IL-10 action did not reduce inflammatory process of dysbiosis [40]. Only one study reported the effect of mechanical treatment on gingivitis control measures by cytokines levels, and the results showed that, although occurs reduction of inflammatory process, the levels of IL-1β, I6 and IL-8 remain high compared to subjects without CP [8]. In conclusion, constipation has a direct detrimental effect on quality of life, which is influenced by antiepileptic drugs intake, type of antiepileptic drugs, and inflammatory signals.

## Data Availability

All relevant data are within the manuscript and its Supporting Information files.

## Acknowledgments

This study was supported by the São Paulo Research Foundation (Fundação de Amparo à Pesquisa do Estado de São Paulo, FAPESP #2017/15160-4.

## Figure Legends

**Supplementary Figure 1:**
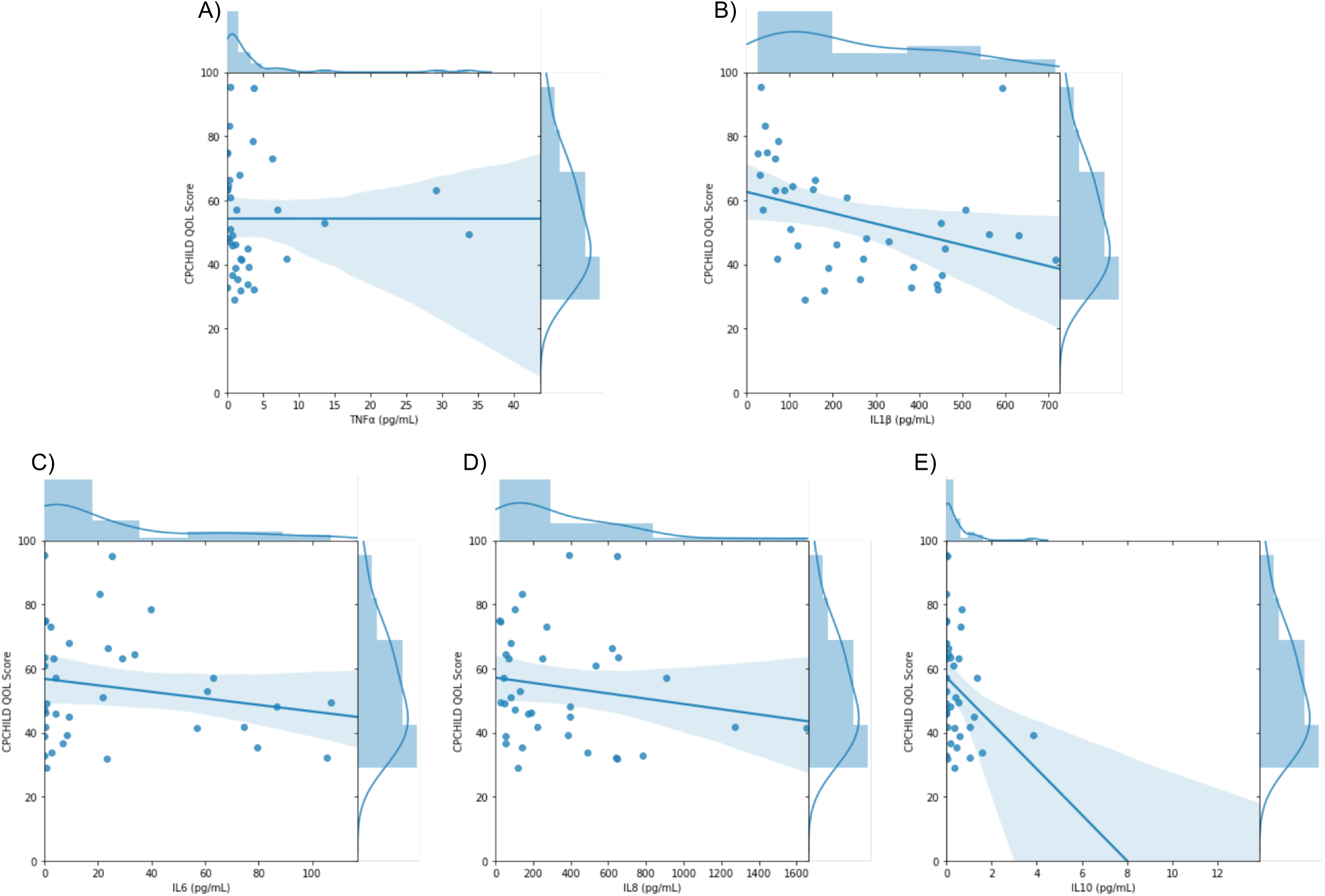
Quality of Life association with specific cytokines. Linear Regressions of collected cytokine levels (TNF-α, IL-1β, IL-6, IL-8, and IL-10 were levels were collected from subjects) and QOL. R- and p-values are displayed, and distribution for each axis is displayed in the opposite margins.

**Supplementary Figure 2:**
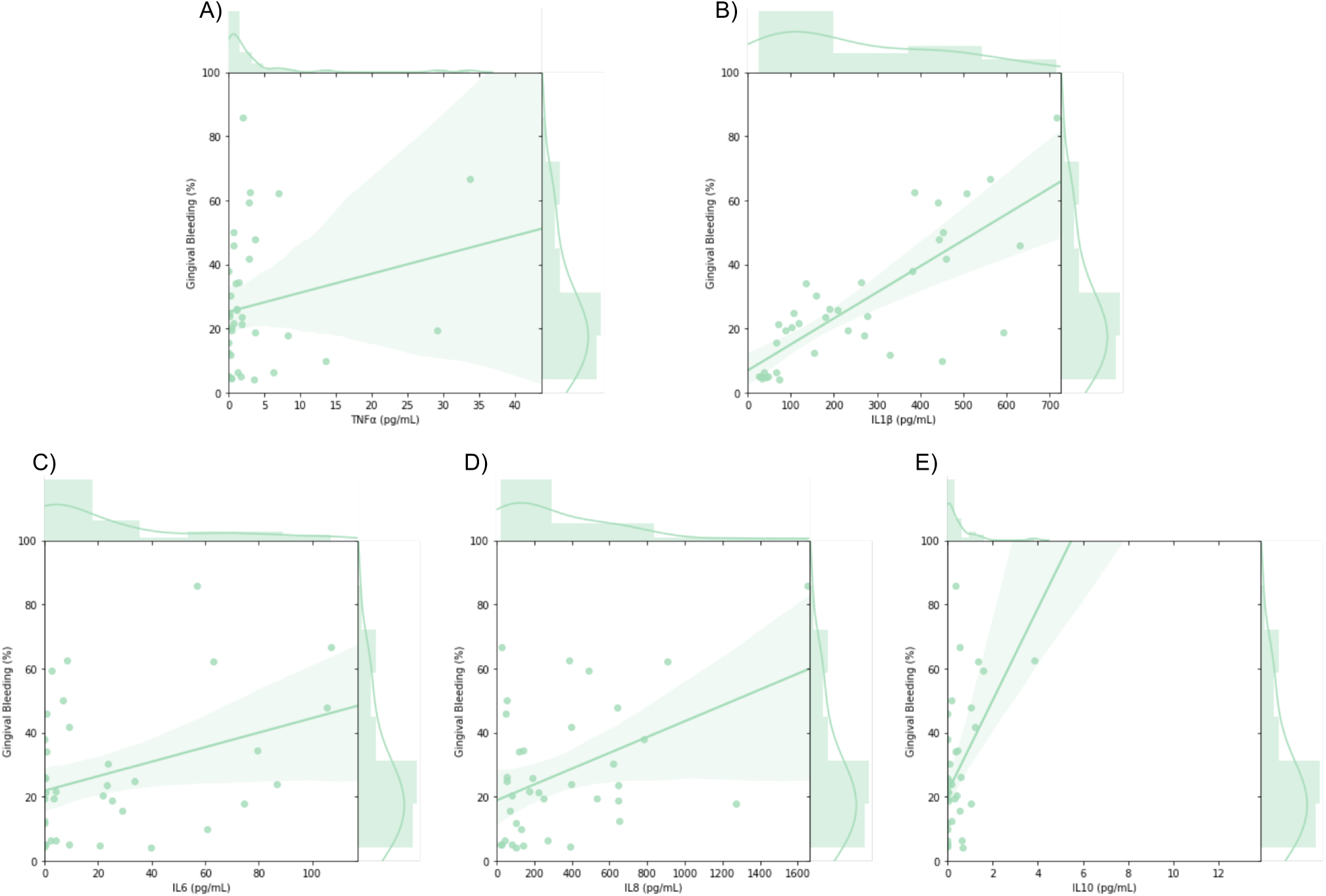
Gingival bleeding association with specific cytokines. Linear Regressions of collected cytokine levels (TNF-α, IL-1β, IL-6, IL-8, and IL-10 were levels were collected from subjects) and gingival bleeding. R- and p-values are displayed, and distribution for each axis is displayed in the opposite margins.

